## Supplementary Information File for "Geographic pair-matching in large-scale cluster randomized trials"

Arnold et al.

### Supplementary Information Figures and Tables

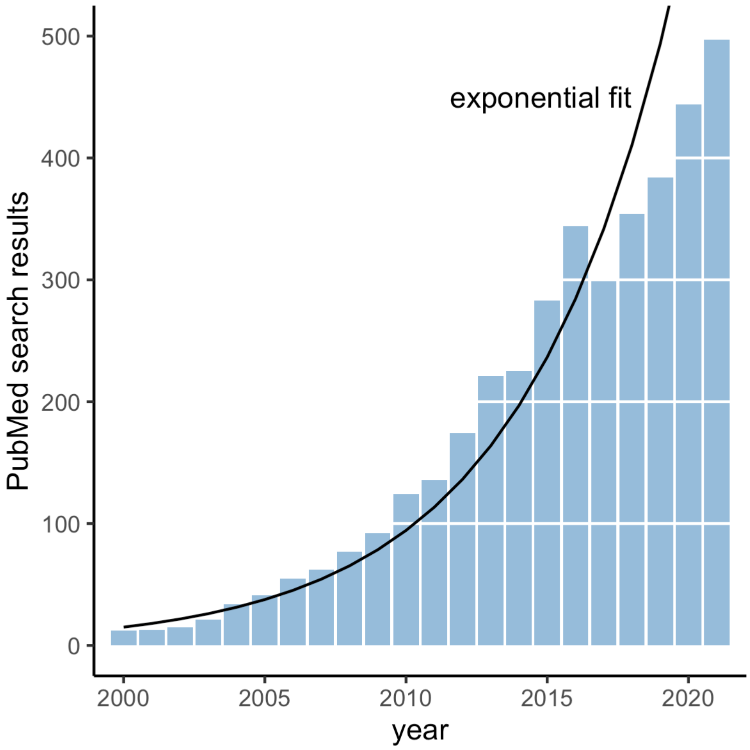

**Supplementary Fig. 1. PubMed search results for cluster randomized trials.** Number of articles indexed on PubMed by year of publication using the query [("cluster randomized trial") OR ("group randomized trial")]. The curve shows predictions from an exponential model fit of the observed counts. Created with notebook https://osf.io/tpksh.

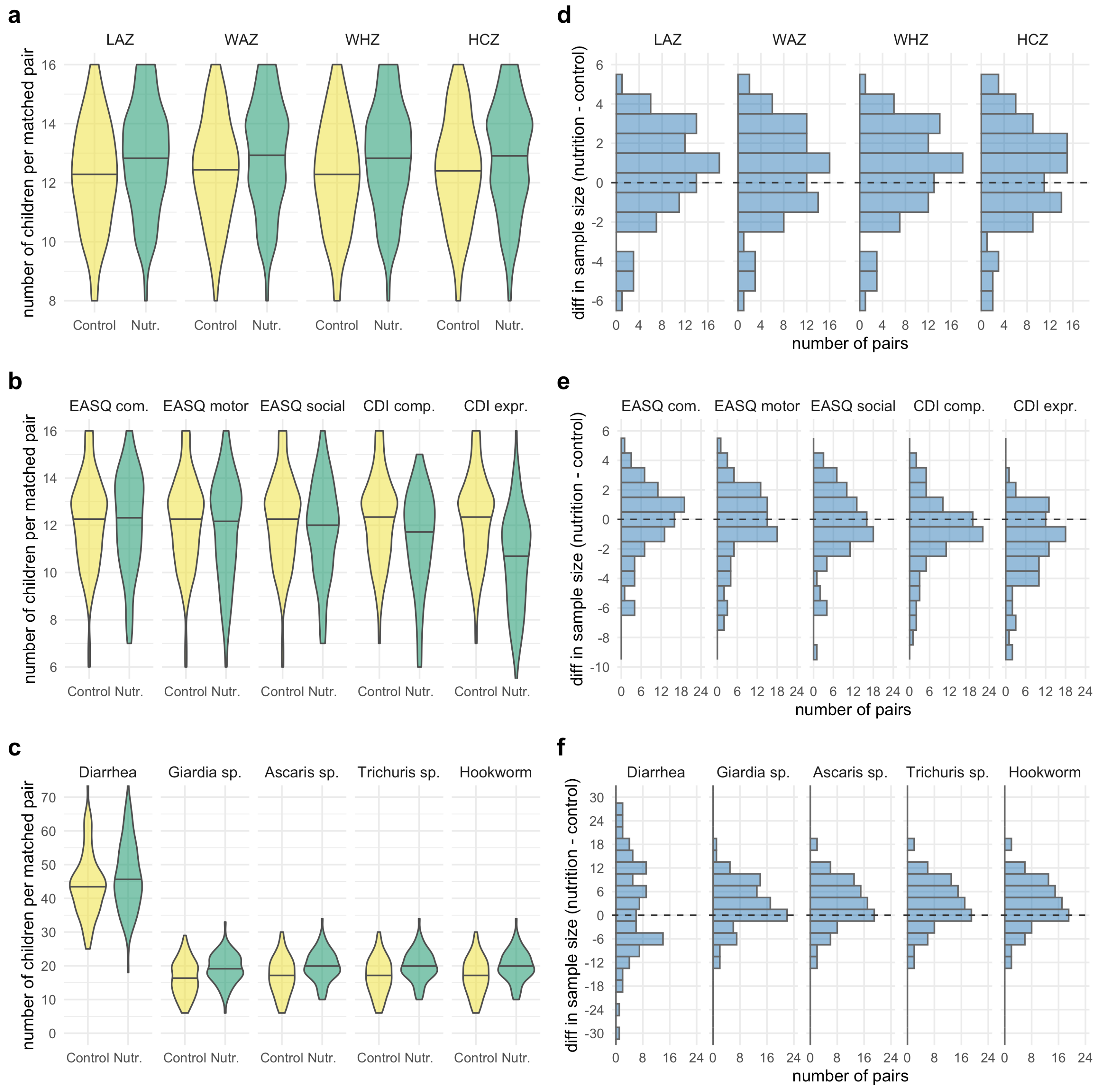

**Supplementary Fig. 2. Distribution of matched pair sample sizes in the Bangladesh trial.** Violin plots summarize the distribution of matched pair sample sizes in the control and nutrition groups for child growth outcomes (**a**), child development outcomes (**b**), and infectious disease outcomes (**c**). Sample sizes were larger for infectious disease outcomes because diarrhea measurements were collected at two timepoints during the trial (at ages 12 and 24 months) and 1-2 older siblings were included from each study compound (Methods). Horizontal lines mark the median of each distribution. Histograms summarize the distribution of differences in sample size between the nutrition and control groups within the 90 matched pairs for child growth outcomes (bin width = 1) (**d**), child development outcomes (bin width = 1) (**e**), and infectious disease outcomes (bin width = 3) (**f**). Supplementary information Table 1 includes sample sizes for each outcome. Created with notebook [https://osf.io/7jhe3](https://osf.io/7jhe3) .

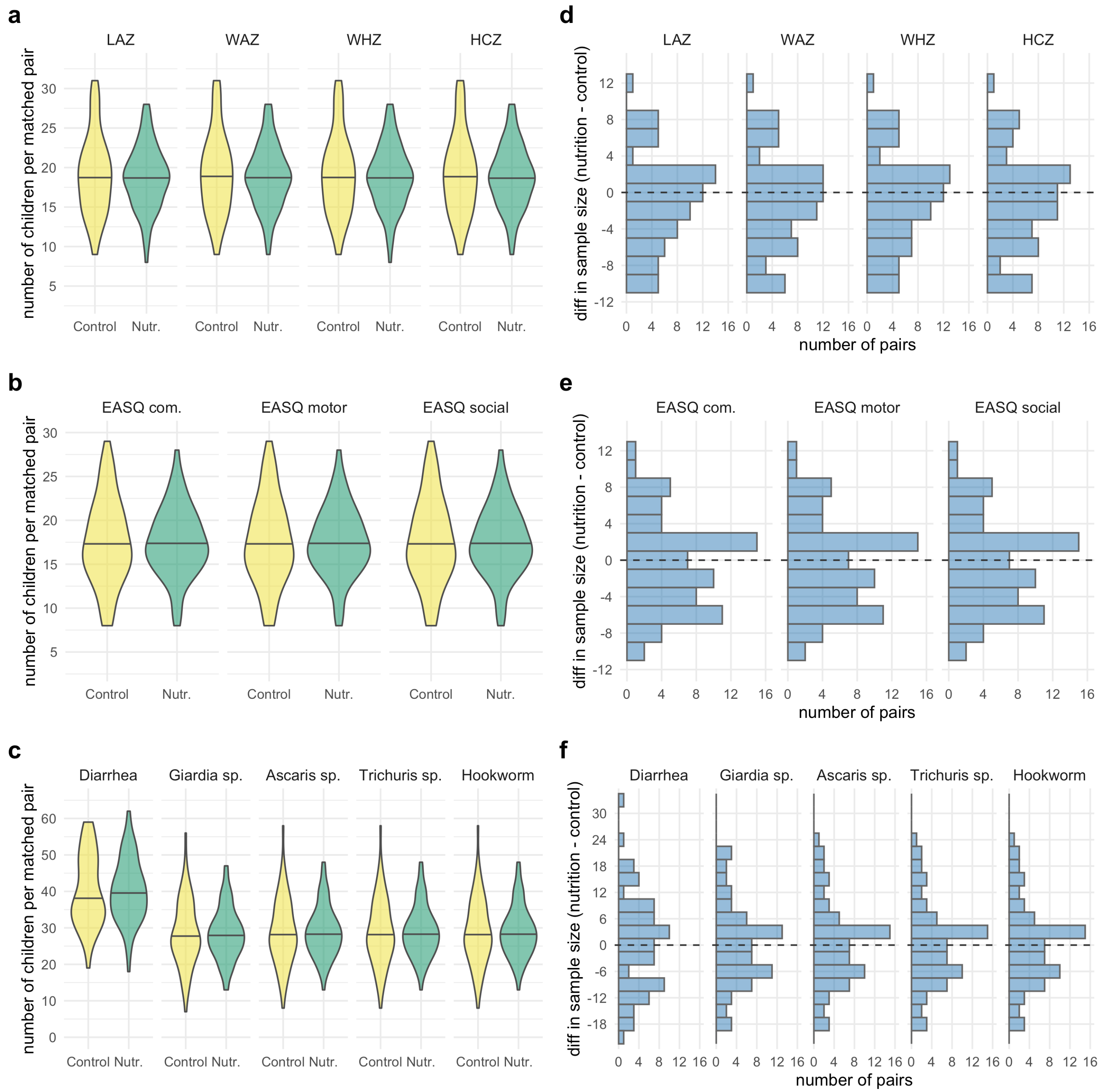

**Supplementary Fig. 3. Distribution of matched pair sample sizes in the Kenya trial.** Violin plots summarize the distribution of matched pair sample sizes in the control and nutrition groups for child growth outcomes (**a**), child development outcomes (**b**), and infectious disease outcomes (**c**). Sample sizes were larger for infectious disease outcomes because diarrhea measurements were collected at two timepoints during the trial (at ages 12 and 24 months) and 1-2 older siblings were included from each study compound (Methods). Horizontal lines mark the median of each distribution. Histograms summarize the distribution of differences in sample size between the nutrition and control groups within the 72 matched pairs for child growth outcomes (bin width = 2) (**d**), child development outcomes (bin width = 2) (**e**), and infectious disease outcomes (bin width = 3) (**f**). Supplementary information Table 2 includes sample sizes for each outcome. Created with notebook https://osf.io/7jhe3 .

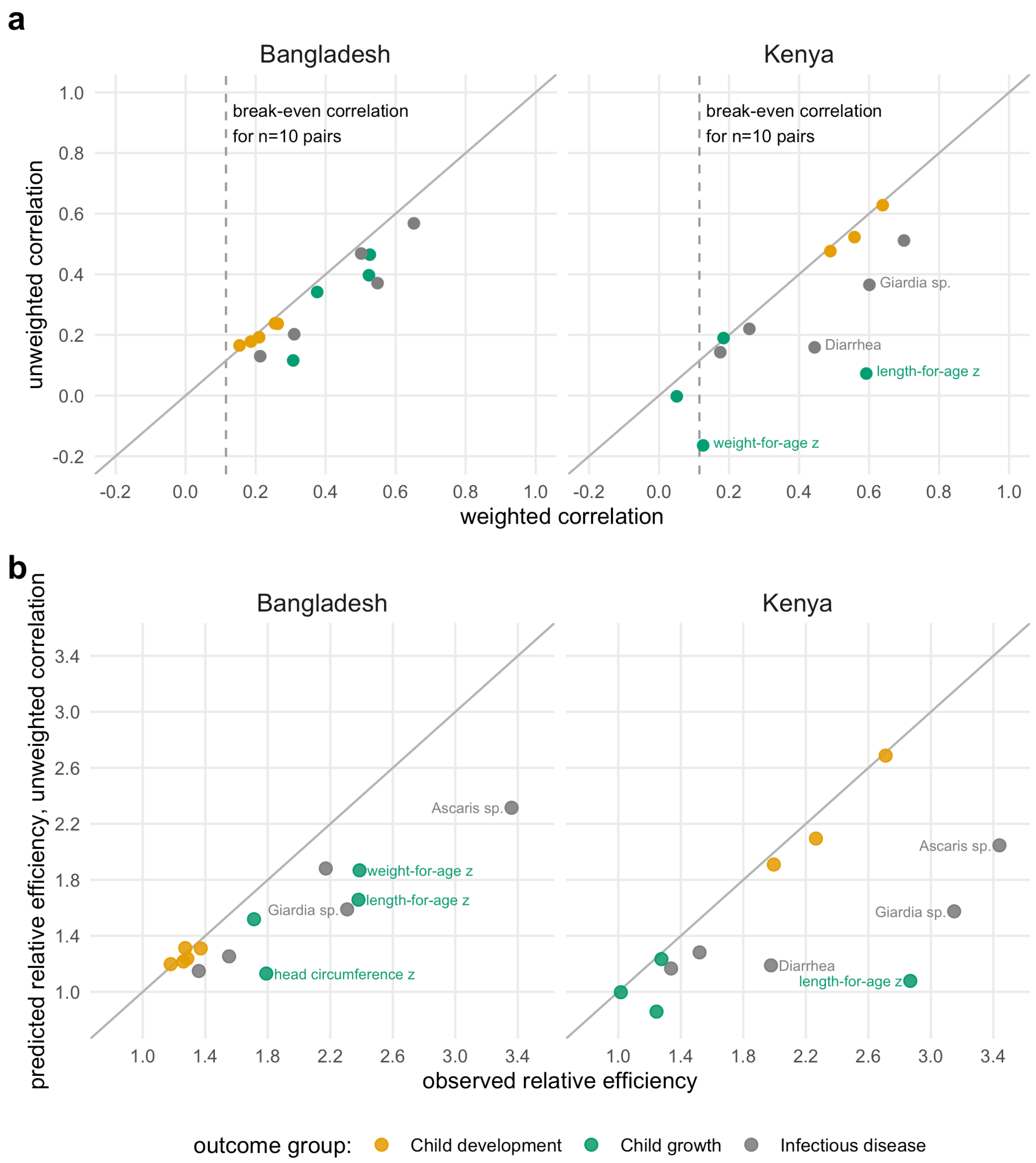

**Supplementary Fig. 4. Different measures of correlation between geographically paired outcomes and relationship with observed relative efficiency.**

**a** Unweighted versus weighted measures of pair-level correlation for 14 child development, child growth, and infectious disease outcomes. Dashed lines show the break-even correlation for a study with 10 matched pairs ($r$ = 0.11). Within-pair outcome correlations higher than the break-even correlation favor a matched design compared with an unmatched design based on statistical efficiency. Solid lines mark the 1:1 axis. Weighted correlations were estimated using pair-level sample sizes as weights. Points are labeled if the weighted correlation is >0.2 higher than the unweighted correlation. **b** Efficiency gains predicted based on the unweighted correlation in panel a versus observed relative efficiency of a the non-parametric, pair-matched estimator compared to an unmatched analysis in the trials. Estimates that differ by >0.4 are labeled. Points that fall below the 1:1 line illustrate empirical efficiency gains that are higher than predicted based on unweighted correlation. Main text Fig 2 includes the relationship for weighted correlation. Communicative Development Inventory (CDI) comprehension and CDI expression were only measured in the Bangladesh trial, hence there are 12 points in the Kenya panel. Created with notebooks https://osf.io/pdver and https://osf.io/d2x3b.

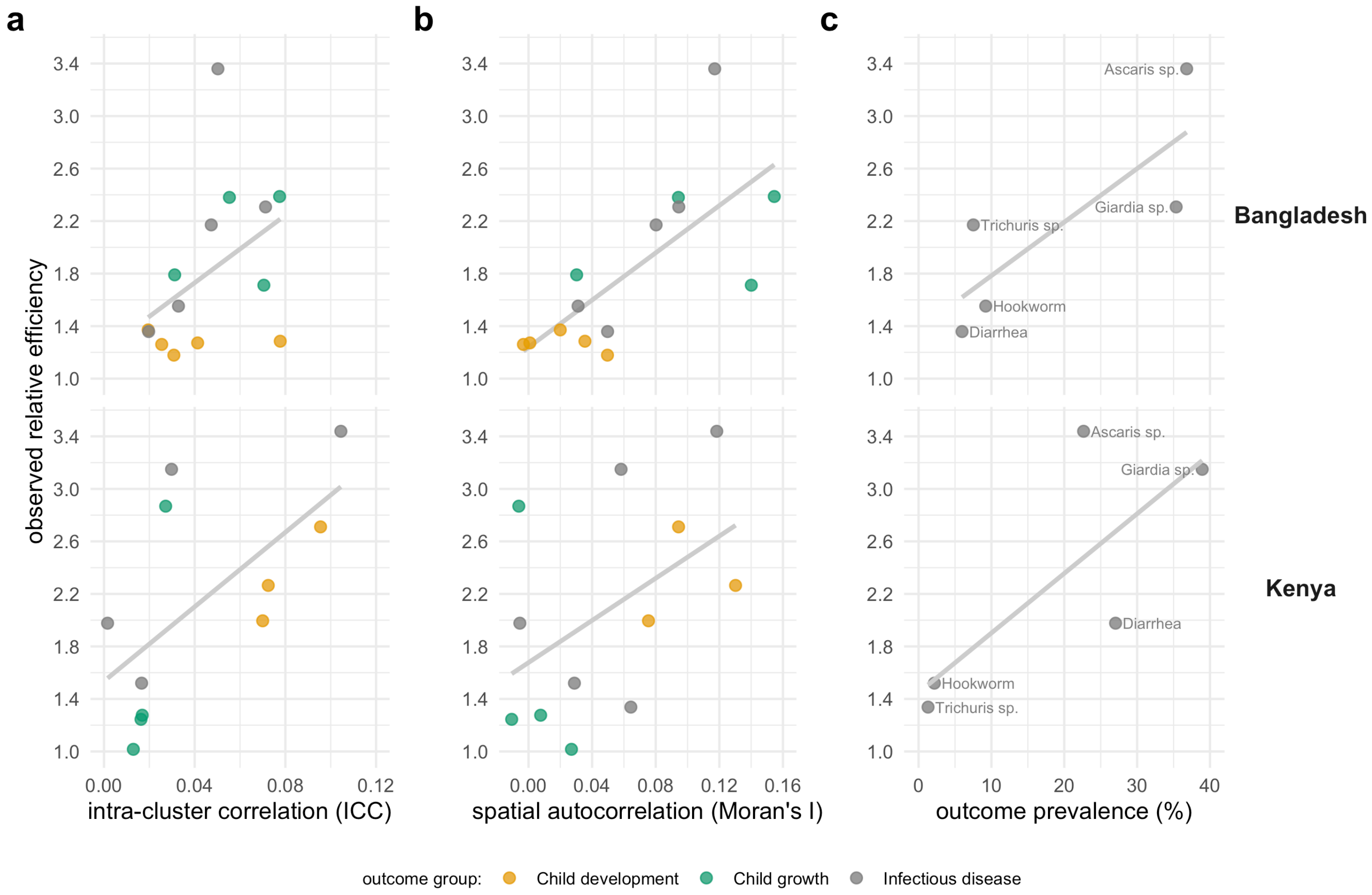

**Supplementary Fig. 5. Relative efficiency of geographic pair-matching compared to an unmatched design as a function of outcome characteristics.**

Panels include 14 children development, child growth, and infectious disease outcomes in the Bangladesh and Kenya WASH Benefits trials**.** Lines in each panel are linear fits to illustrate trends. **a** Observed relative efficiency of geographic pair-matching versus outcome intra-cluster correlation (ICC). **b** Observed relative efficiency of geographic pair-matching versus outcome spatial autocorrelation as measured by Moran's I. **c** Observed relative efficiency of geographic pair-matching versus outcome prevalence for infectious disease outcomes (labeled). ICC, Moran's I, and outcome prevalence (x-axes) were estimated using control clusters only. The ICC for *Trichuris* sp. could not be estimated consistently in Kenya due to very low prevalence. Communicative Development Inventory (CDI) comprehension and CDI expression were only measured in the Bangladesh trial. Created with notebooks https://osf.io/pdver , https://osf.io/rbp38, https://osf.io/827wz, and https://osf.io/d2x3b.

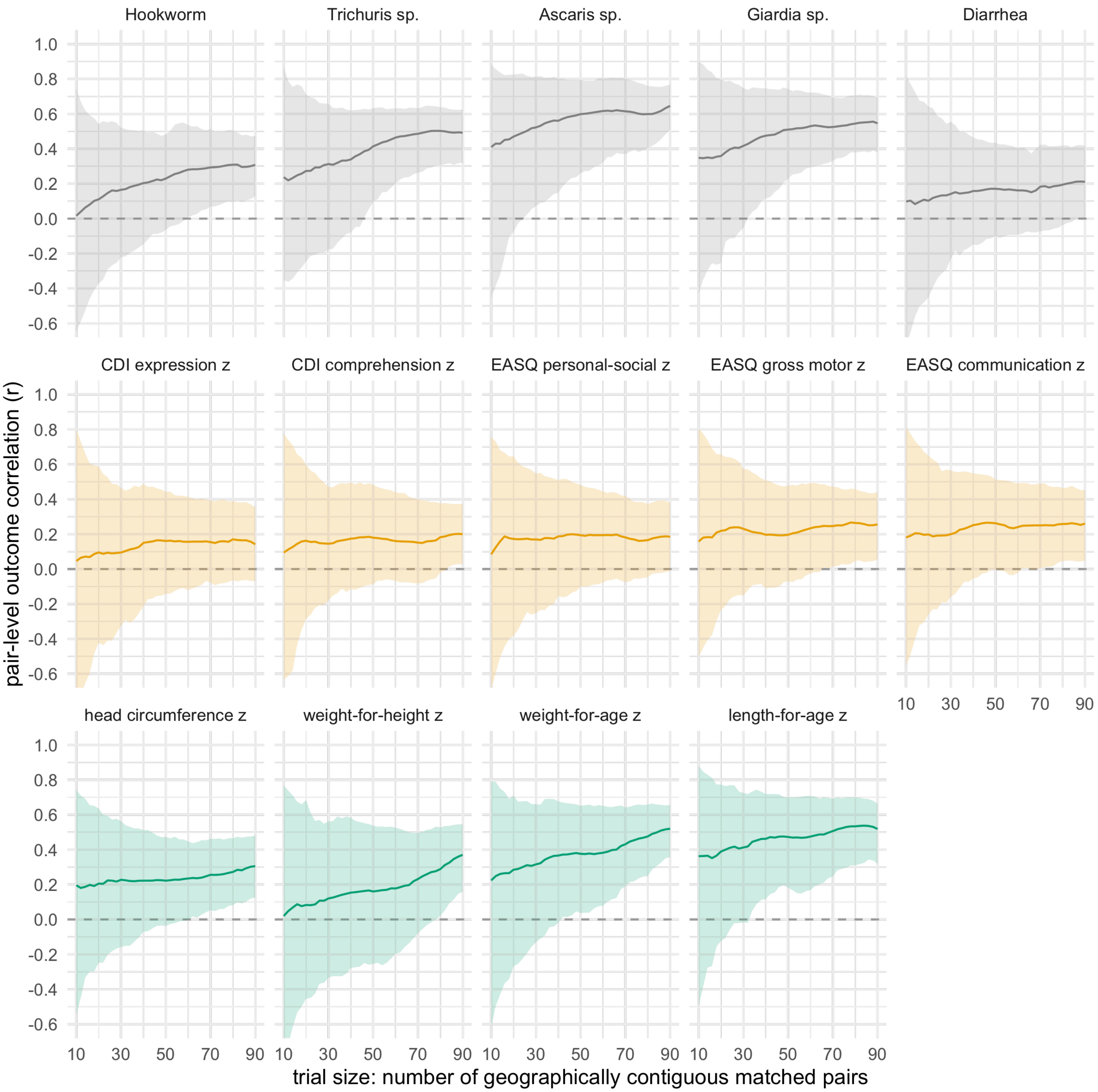

**Supplementary Fig. 6. Pair-level outcome correlation in the Bangladesh trial by outcome and by number of geographically contiguous matched pairs.**
Lines represent means and shaded regions 95% bootstrapped confidence intervals for the cluster-level outcome correlation between matched pairs at different sample sizes. For each sample size between $n$ = 10, 12, …, 90 pairs, an index pair was randomly selected from the overall trial and $n$–1 geographically proximate pairs were sampled with replacement. This modified bootstrap was repeated 1,000 times at each sample size. Created with notebook https://osf.io/n276c .

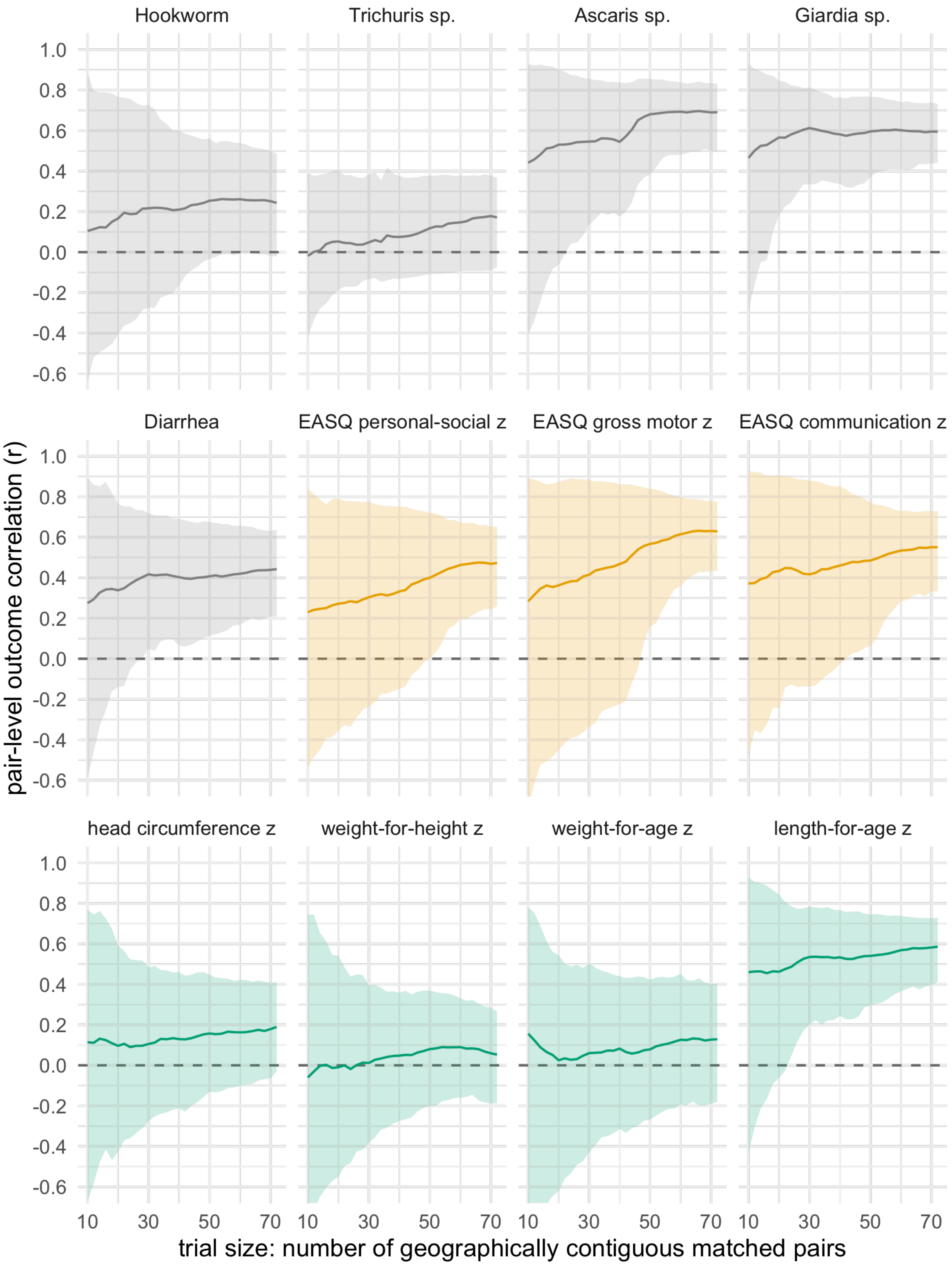

**Supplementary Fig. 7. Pair-level outcome correlation in the Kenya trial by outcome and by number of geographically contiguous matched pairs.**

Lines represent means and shaded regions 95% bootstrapped confidence intervals for the cluster-level outcome correlation between matched pairs at different sample sizes. For each sample size between *n* = 10, 12, …, 72 pairs, an index pair was randomly selected from the overall trial and *n*–1 geographically proximate pairs were sampled with replacement. This modified bootstrap was repeated 1,000 times at each sample size. Created with notebook https://osf.io/n276c .

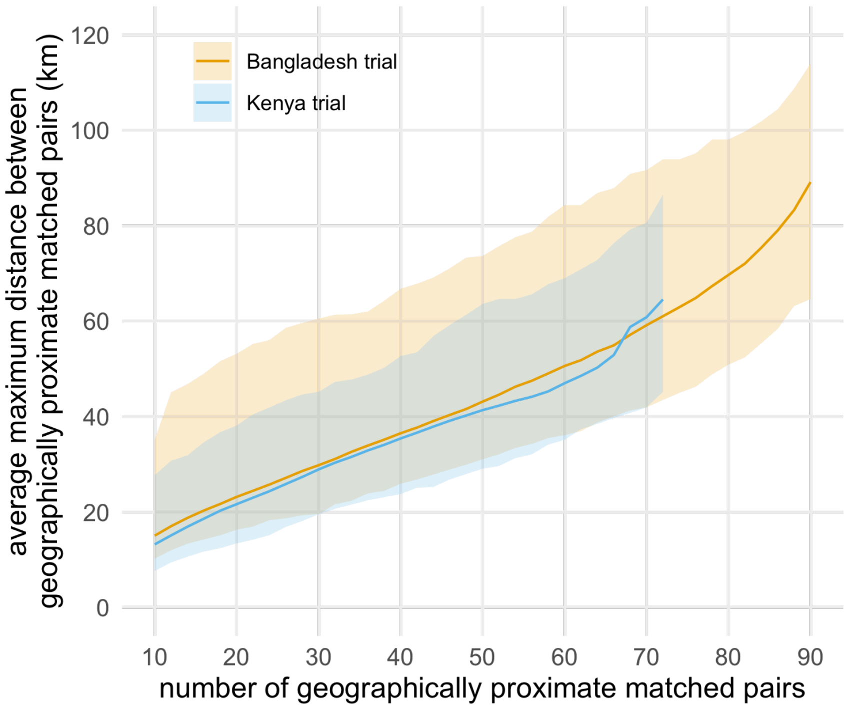

**Supplementary Fig. 8. Geographic footprint of the trials across a range of geographically proximate matched pairs.**

Lines represent means and shaded regions 95% bootstrapped confidence intervals for the cluster-level outcome correlation between matched pairs at different sample sizes. For each sample size between *n* = 10, 12, …, to the maximum number of pairs in each trial (90 in Bangladesh, 72 in Kenya), an index pair was randomly selected from the overall trial and *n*–1 geographically proximate pairs were sampled with replacement. This modified bootstrap was repeated 1,000 times at each sample size. Created with notebook https://osf.io/n276c .

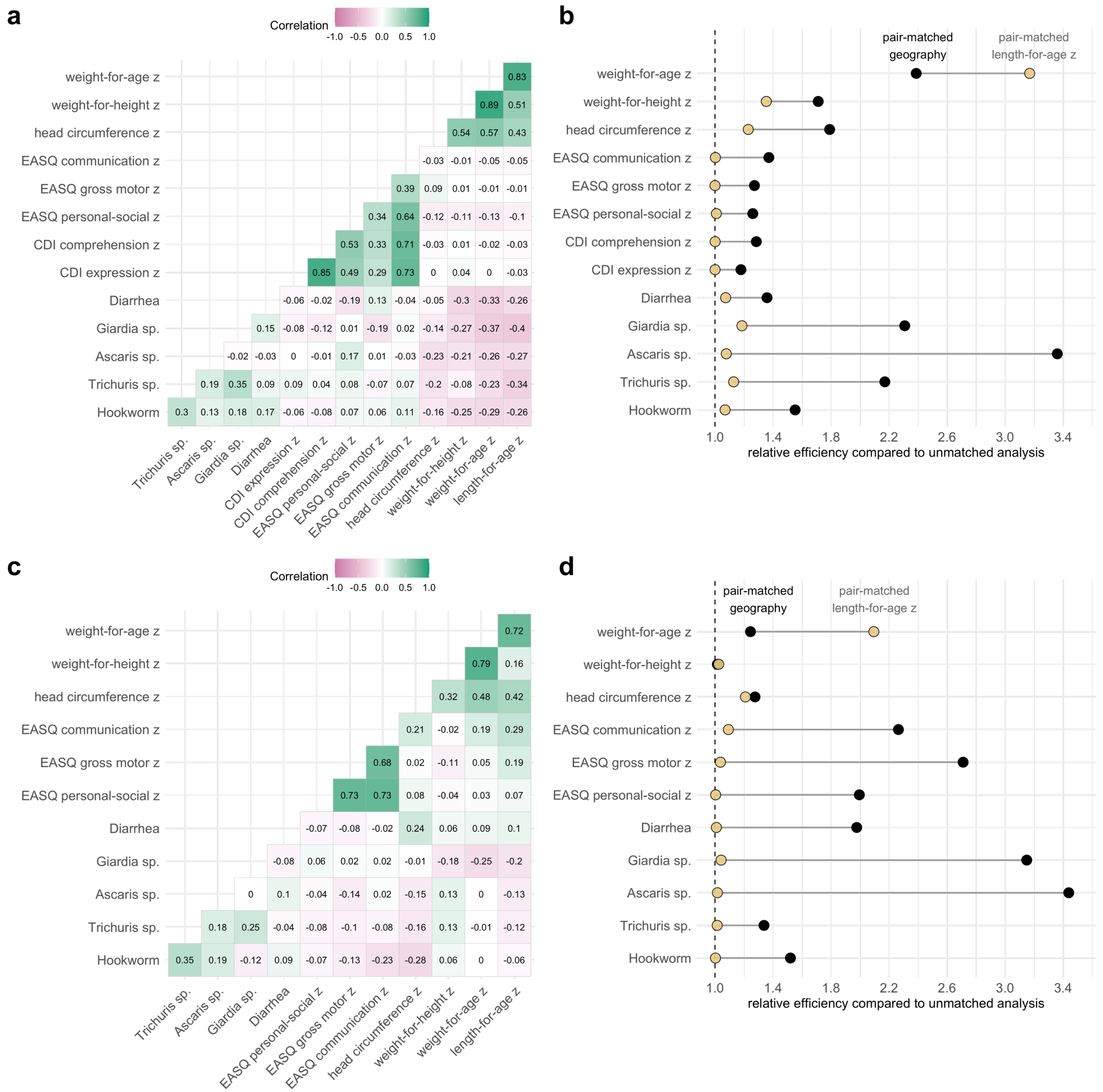

**Supplementary Fig. 9. Correlation between outcomes in and corresponding estimates of relative efficiency from pair-matching on length-for-age z, a primary outcome**
**a** Pair-level outcome correlations in the 90 control means for the WASH Benefits Bangladesh trial. **b** Approximate estimates of relative efficiency of pair-matching on length-for-age z derived from outcome correlations in the rightmost column of panel a (Methods) and compared with observed relative efficiency of geographic pair matching in the Bangladesh trial. **c** Pair-level outcome correlations in the 72 control means for the WASH Benefits Kenya trial. **d** Approximate estimates of relative efficiency of pair-matching on length-for-age z derived from outcome correlations in the rightmost column of panel c (Methods) and compared with observed relative efficiency of geographic pair-matching in the Kenya trial. Created with notebook [https://osf.io/nf3j4](https://osf.io/nf3j4) .

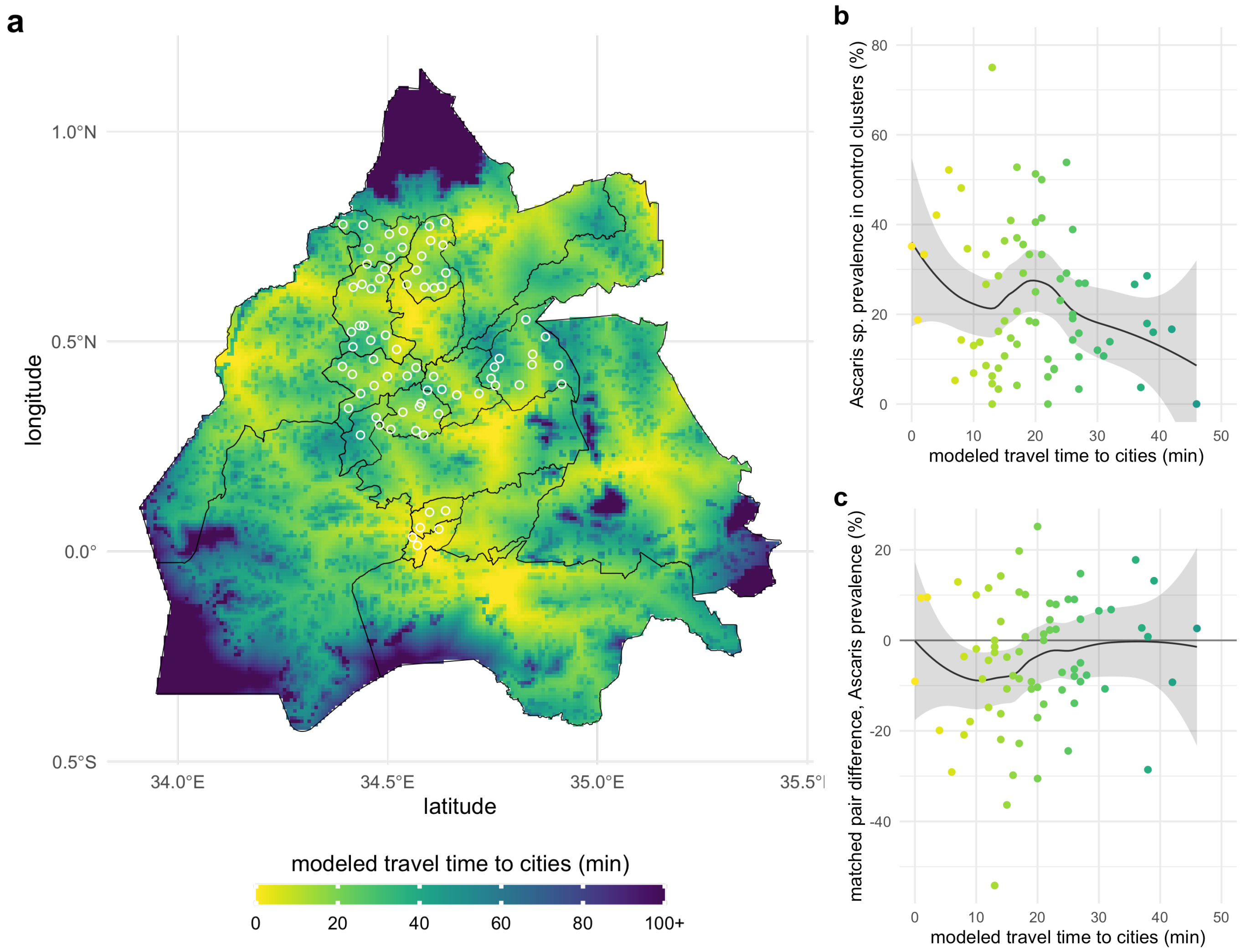

**Supplementary Fig. 10. Heterogeneity in the effect of nutrition on *Ascaris* sp. prevalence by travel time to cities, Kenya.**

**a** Modeled travel time to cities in minutes and the 72 WASH Benefits Kenya matched pair centroids (white circles). Black lines mark sub-counties. **b** *Ascaris* sp. infection prevalence in control clusters by travel time to cities. **c** Matched pair differences in *Ascaris* sp. infection prevalence (nutrition – control) by travel time to cities. In panels b and c, points are colored by the surface in panel a, the line represents a non-parametric locally weighted regression fit, and the shaded band its approximate pointwise 95% confidence interval. Created with notebook https://osf.io/fmgex .

**Supplementary Table 1. Bangladesh trial outcome summary.**

Sample sizes (n) differ by outcome within child growth and child development outcome groups based on missingness, and differ within infectious disease outcomes based on different measurement strategies. Outcomes were measured at the final visit when birth cohort children were approximately 24 months old except for diarrhea, which also included measurements when birth children were approximately 12 months old. Infectious disease outcome measurements included older siblings (Methods). Created with notebook https://osf.io/7jhe3 .

| | | Control | | | Nutrition | | | | | |
|---|---|---|---|---|---|---|---|---|---|---|
| Outcome | Median age y (range) | n | mean | sd | n | mean | sd | ATE (95% CI)* | ICC (95% CI)† | Moran's I |
| **Child growth** | | | | | | | | | | |
| length-for-age z | 1.9 (1.4, 3.1) | 1,103 | -1.79 | 1.01 | 1,158 | -1.60 | 1.03 | 0.18 (0.10, 0.26) | 0.06 (0.02, 0.09) | 0.09 |
| weight-for-age z | 1.9 (1.4, 3.1) | 1,121 | -1.54 | 1.00 | 1,165 | -1.36 | 1.03 | 0.18 (0.09, 0.26) | 0.08 (0.04, 0.13) | 0.15 |
| weight-for-height z | 1.9 (1.4, 3.1) | 1,104 | -0.88 | 0.93 | 1,158 | -0.75 | 0.97 | 0.12 (0.03, 0.20) | 0.07 (0.03, 0.11) | 0.14 |
| head circumference z | 1.9 (1.4, 3.1) | 1,118 | -1.61 | 0.94 | 1,160 | -1.48 | 0.92 | 0.13 (0.05, 0.21) | 0.03 (0.00, 0.06) | 0.03 |
| **Child development** | | | | | | | | | | |
| EASQ communication z | 2.1 (1.7, 2.5) | 1,099 | 0.00 | 1.00 | 1,094 | 0.22 | 0.88 | 0.23 (0.15, 0.30) | 0.02 (0.00, 0.05) | 0.02 |
| EASQ gross motor z | 2.1 (1.7, 2.5) | 1,099 | 0.00 | 1.00 | 1,074 | 0.16 | 0.94 | 0.17 (0.08, 0.25) | 0.04 (0.01, 0.08) | 0.00 |
| EASQ personal-social z | 2.1 (1.7, 2.5) | 1,099 | 0.00 | 1.00 | 1,066 | 0.28 | 0.98 | 0.28 (0.20, 0.37) | 0.03 (0.00, 0.06) | 0.00 |
| CDI comprehension z | 2.1 (1.7, 2.5) | 1,106 | 0.00 | 1.00 | 1,029 | 0.22 | 0.83 | 0.22 (0.13, 0.31) | 0.08 (0.03, 0.12) | 0.04 |
| CDI expression z | 2.1 (1.7, 2.5) | 1,106 | 0.00 | 1.00 | 945 | 0.19 | 0.89 | 0.19 (0.10, 0.27) | 0.03 (0.00, 0.06) | 0.05 |
| **Infectious disease** | | | | | | | | | | |
| Diarrhea | 1.8 (0.0, 5.5) | 4,022 | 0.06 | 0.24 | 4,226 | 0.04 | 0.19 | -0.02 (-0.03, -0.01) | 0.02 (0.01, 0.03) | 0.05 |
| Giardia sp. | 2.7 (2.1, 12.0) | 1,468 | 0.35 | 0.48 | 1,724 | 0.31 | 0.46 | -0.04 (-0.08, -0.01) | 0.07 (0.03, 0.11) | 0.09 |
| Ascaris sp. | 2.7 (2.1, 12.0) | 1,530 | 0.37 | 0.48 | 1,796 | 0.37 | 0.48 | 0.01 (-0.03, 0.04) | 0.05 (0.02, 0.08) | 0.12 |
| Trichuris sp. | 2.7 (2.1, 12.0) | 1,530 | 0.08 | 0.26 | 1,796 | 0.08 | 0.27 | 0.02 (-0.01, 0.04) | 0.05 (0.02, 0.08) | 0.08 |
| Hookworm | 2.7 (2.1, 12.0) | 1,530 | 0.09 | 0.29 | 1,796 | 0.08 | 0.27 | -0.01 (-0.03, 0.01) | 0.03 (0.01, 0.06) | 0.03 |

* ATE: Average treatment effect of difference in means (Nutrition minus Control), with 95% confidence interval reported for the non-parametric, pair-matched estimator. For infectious disease outcomes, this is the difference in the proportion positive.

† ICC: Intra-cluster correlation coefficient among outcomes measured in the control group, with 95% confidence interval estimated through bootstrap resampling geographic pairs with replacement (1,000 iterations).

**Supplementary Table 2. Kenya trial outcome summary.**
Sample sizes (n) differ by outcome within child growth and child development outcome groups based on missingness, and differ within infectious disease outcomes based on different measurement strategies. Outcomes were measured at the final visit when birth cohort children were approximately 24 months old except for diarrhea, which also included measurements when birth children were approximately 12 months old. Infectious disease outcome measurements included older siblings (Methods). Created with notebook https://osf.io/7jhe3 .

| | | Control | | | Nutrition | | | | | |
|---|---|---|---|---|---|---|---|---|---|---|
| **Outcome** | **Median age y (range)** | **n** | **mean** | **sd** | **n** | **mean** | **sd** | **ATE (95% CI)*** | **ICC (95% CI)†** | **Moran's I** |
| **Child growth** | | | | | | | | | | |
| length-for-age z | 2.1 (1.4, 2.6) | 1,378 | -1.55 | 1.11 | 1,350 | -1.42 | 1.08 | 0.14 (0.05, 0.23) | 0.03 (0.01, 0.05) | -0.01 |
| weight-for-age z | 2.1 (1.4, 2.6) | 1,390 | -0.73 | 1.01 | 1,357 | -0.62 | 0.98 | 0.11 (0.03, 0.20) | 0.02 (0.00, 0.04) | -0.01 |
| weight-for-height z | 2.1 (1.4, 2.6) | 1,379 | 0.11 | 0.95 | 1,352 | 0.16 | 0.91 | 0.05 (-0.03, 0.13) | 0.01 (0.00, 0.04) | 0.03 |
| head circumference z | 2.1 (1.4, 2.6) | 1,387 | -0.28 | 1.00 | 1,352 | -0.23 | 1.00 | 0.05 (-0.03, 0.13) | 0.02 (0.00, 0.04) | 0.01 |
| **Child development** | | | | | | | | | | |
| EASQ communication z | 2.1 (1.6, 2.5) | 1,264 | 0.00 | 1.01 | 1,264 | 0.00 | 0.99 | 0.02 (-0.06, 0.10) | 0.07 (0.04, 0.11) | 0.13 |
| EASQ gross motor z | 2.1 (1.6, 2.5) | 1,264 | 0.01 | 1.00 | 1,264 | -0.01 | 0.96 | -0.00 (-0.08, 0.07) | 0.10 (0.05, 0.15) | 0.09 |
| EASQ personal-social z | 2.1 (1.6, 2.5) | 1,264 | 0.01 | 1.01 | 1,264 | 0.02 | 0.97 | 0.01 (-0.07, 0.10) | 0.07 (0.03, 0.11) | 0.08 |
| **Infectious disease** | | | | | | | | | | |
| Diarrhea | 1.8 (0.0, 4.7) | 2,881 | 0.27 | 0.44 | 2,895 | 0.27 | 0.44 | 0.00 (-0.02, 0.03) | 0.00 (0.00, 0.01) | -0.01 |
| Giardia sp. | 3.4 (2.0, 15.0) | 2,050 | 0.39 | 0.49 | 2,045 | 0.40 | 0.49 | 0.01 (-0.02, 0.04) | 0.03 (0.01, 0.05) | 0.06 |
| Ascaris sp. | 3.4 (2.0, 15.0) | 2,075 | 0.23 | 0.42 | 2,085 | 0.19 | 0.39 | -0.04 (-0.07, -0.01) | 0.10 (0.06, 0.15) | 0.12 |
| Trichuris sp. | 3.4 (2.0, 15.0) | 2,075 | 0.01 | 0.11 | 2,085 | 0.01 | 0.08 | -0.01 (-0.02, 0.00) | ‡ | 0.06 |
| Hookworm | 3.4 (2.0, 15.0) | 2,075 | 0.02 | 0.15 | 2,085 | 0.03 | 0.18 | 0.01 (-0.00, 0.02) | 0.02 (0.00, 0.04) | 0.03 |

* ATE: Average treatment effect of difference in means (Nutrition minus Control), with 95% confidence interval reported for the non-parametric, pair-matched estimator. For infectious disease outcomes, this is the difference in the proportion positive.

† ICC: Intra-cluster correlation coefficient among outcomes measured in the control group, with 95% confidence interval estimated through bootstrap resampling geographic pairs with replacement (1,000 iterations).

‡ The mixed effects model did not converge for Trichuris sp. infection in Kenya, so no estimate of the ICC was possible.

§ MacArthur-Bates Communicative Development Inventory (CDI) measures were not collected in the Kenya trial.

**Supplementary Table 3. Pair-level correlation and relative efficiency for outcomes in the Bangladesh and Kenya trials.**

Outcome correlation between control and nutrition groups within matched pairs and corresponding estimates of relative efficiency of geographic pair-matching compared to an unmatched design (Methods). Created with notebook https://osf.io/7jhe3 .

| | Pair-wise correlation (95% CI)* | | Relative Efficiency (95% CI)† | |
|---|---|---|---|---|
| **Outcome** | **Bangladesh** | **Kenya** | **Bangladesh** | **Kenya** |
| **Child growth** | | | | |
| length-for-age z | 0.52 (0.34, 0.67) | 0.59 (0.42, 0.73) | 2.1 (1.5, 3.0) | 2.5 (1.7, 3.7) |
| weight-for-age z | 0.53 (0.35, 0.66) | 0.13 (-0.17, 0.42) | 2.1 (1.5, 3.0) | 1.1 (0.9, 1.7) |
| weight-for-height z | 0.38 (0.16, 0.54) | 0.05 (-0.17, 0.27) | 1.6 (1.2, 2.2) | 1.1 (0.9, 1.4) |
| head circumference z | 0.31 (0.12, 0.46) | 0.18 (-0.05, 0.40) | 1.4 (1.1, 1.9) | 1.2 (1.0, 1.7) |
| **Child development** | | | | |
| EASQ communication z | 0.26 (0.07, 0.46) | 0.56 (0.34, 0.71) | 1.4 (1.1, 1.8) | 2.3 (1.5, 3.4) |
| EASQ gross motor z | 0.26 (0.07, 0.44) | 0.64 (0.45, 0.77) | 1.3 (1.1, 1.8) | 2.8 (1.8, 4.4) |
| EASQ personal-social z | 0.19 (-0.02, 0.39) | 0.49 (0.24, 0.64) | 1.2 (1.0, 1.6) | 2.0 (1.3, 2.8) |
| CDI comprehension z | 0.21 (0.02, 0.39) | ‡ | 1.3 (1.0, 1.6) | ‡ |
| CDI expression z | 0.15 (-0.07, 0.36) | ‡ | 1.2 (0.9, 1.6) | ‡ |
| **Infectious disease** | | | | |
| Diarrhea | 0.21 (-0.01, 0.45) | 0.44 (0.23, 0.62) | 1.3 (1.0, 1.8) | 1.8 (1.3, 2.6) |
| Giardia sp. | 0.55 (0.38, 0.68) | 0.60 (0.44, 0.73) | 2.2 (1.6, 3.1) | 2.5 (1.8, 3.7) |
| Ascaris sp. | 0.65 (0.50, 0.75) | 0.70 (0.50, 0.83) | 2.9 (2.0, 4.0) | 3.3 (2.0, 5.7) |
| Trichuris sp. | 0.50 (0.31, 0.62) | 0.17 (-0.07, 0.37) | 2.0 (1.5, 2.6) | 1.2 (0.9, 1.6) |
| Hookworm | 0.31 (0.13, 0.48) | 0.26 (-0.01, 0.48) | 1.4 (1.1, 1.9) | 1.3 (1.0, 1.9) |

* Weighted correlation between geographically paired, cluster-level means. 95% confidence interval estimated using a non-parametric bootstrap of pairs.

† Relative efficiency of geographic pair-matching compared to an unmatched design, defined as 1/(1-r), where r is the weighted correlation between paired outcomes. 95% confidence interval estimated using a non-parametric bootstrap of pairs.

‡ MacArthur-Bates Communicative Development Inventory (CDI) measures were not collected in the Kenya trial.
